# Structured simulation-based training with frugal ophthalmic instruments rapidly equips non-ophthalmologists to identify malarial retinopathy

**DOI:** 10.1101/2025.01.23.24318545

**Authors:** Kyle J Wilson, Obaid Kousha, Harold Nkume, Alice M Liomba, Nicholas AV Beare, Andrew Blaikie

## Abstract

**Background:** Malaria, particularly cerebral malaria, poses a significant health threat, especially for children in sub-Saharan Africa. Identification of malarial retinopathy (MR) has diagnostic value in cerebral malaria but can be impractical due to lack of availability of trained and equipped ophthalmic specialists. Recent developments in low-cost simulation tools and ophthalmoscopes offer the potential to train non-ophthalmologists to offer fundus assessments in low-resource settings.

**Methods:** Twenty non-ophthalmologist healthcare providers in Malawi attended a training program focused on identification of MR using direct and binocular indirect ophthalmoscopy. The curriculum included didactic sessions, hands-on practice with simulation eyes, and assessment of skill acquisition with immediate feedback. Pre- and post-training confidence surveys were conducted.

**Results:** Participants demonstrated significant improvement in identification of malarial retinopathy after completing the training, with a mean accuracy of 83%. Clinician confidence levels increased across all domains of retinal examination covered in the course.

**Conclusion:** Structured simulation-based training using frugal ophthalmoscopes can equip non-ophthalmologists with skills to identify MR in simulation eyes. Future work should focus on long-term skill retention and validation in a real-world clinical setting.

## Background

Malaria remains a significant public health concern globally, being responsible for more than 600,000 deaths in 2022.^1^ Over 70% of these deaths occur in children in the African region. Within the spectrum of disease caused by malaria infection, cerebral malaria (CM) stands out as the deadliest.^2^ It is characterised by a Blantyre Coma Score ≤ 2 and peripheral parasitaemia. However, in areas with a high burden of asymptomatic peripheral parasitaemia discerning the aetiology of paediatric coma can be challenging, as other diseases may contribute to similar clinical presentations.^3^

Malarial retinopathy (MR) has been shown to have diagnostic and prognostic value in CM.^3,4^ Examination of the fundus for signs of MR is best achieved using binocular indirect ophthalmoscopy (BIO). Where BIO is not available a compromised but adequate examination can be achieved with a direct ophthalmoscope, albeit with a much smaller field of view.^5^ however Access to trained and equipped eye care specialists in paediatric wards and out of hours is, typically impractical in resource-limited regions.^6^

The low-cost Arclight direct ophthalmoscope (Figure 1a) is now established as a quality alternative to expensive traditional devices.^7,8^ More recently, the low-cost Arclight BIO (Figure 1b) has been developed. It has been shown to perform as well as a more expensive traditional BIO, making it suitable for widespread use in low-and-middle-income countries.^9^ Pilot data (Kousha & Blaikie, unpublished) suggests that ophthalmology-naïve health care workers can quickly acquire proficiency with BIO, subject to structured training in a specific-disease area. MR is an ideal target for such training given that: (1) it has diagnostic utility in a disease that is both serious and common; (2) patients with cerebral malaria (usually children in sub-Saharan Africa) are routinely seen by non-ophthalmologists for their clinical care; (3) access to a specialist eye care professional is rarely available.

**Figure 1.**
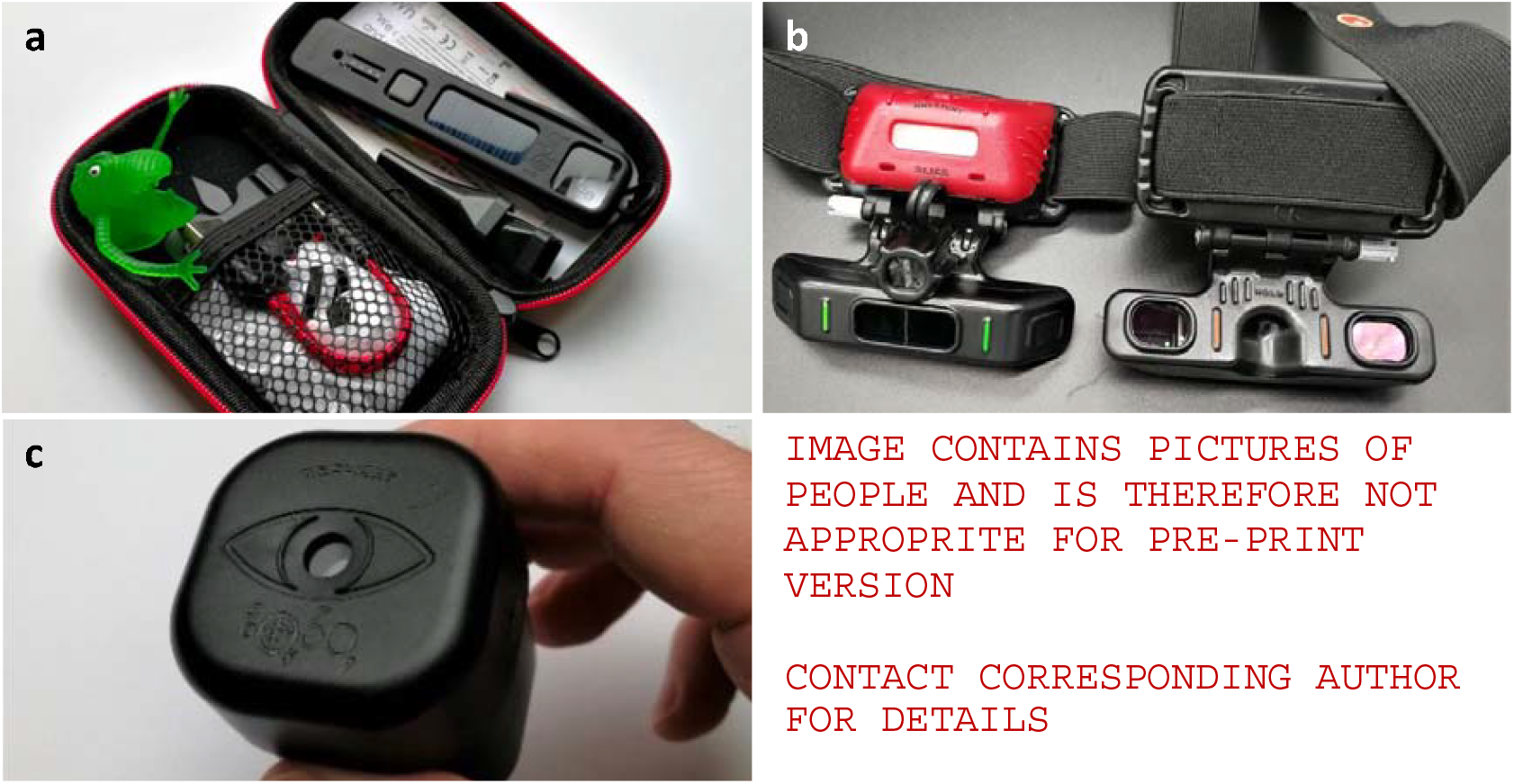
a) The Arclight direct ophthalmoscope. b) The Arclight binocular indirect ophthalmoscope. c) Low-cost simulation eyes have high-fidelity printed retinas and a plastic lens to mimic the optical power of the human eye. promotional image of the Arclight binocular indirect ophthalmoscope in use. All images provided by the Arclight Project and reproduced with permission.

We designed and delivered a training programme in the identification of MR for 20 participants using simulation eyes (Figure 1c 1d). Here, we present the results of an audit of the outcomes of a ‘Fundoscopy for Cerebral Malaria’ training course for a mixed group of non-ophthalmologists in a malaria-endemic area.

## Methods

### Participants

We designed and delivered a training course in ‘Fundoscopy for Cerebral Malaria’ to 20 participants from Queen Elizabeth Central Hospital in Blantyre, Malawi. All participants were either paediatricians or family medicine doctors (55%) or optometrists (45%). None had experience of using a BIO in a clinical environment, though optometrists had been briefly orientated to BIO during their studies.

### Training structure

The training programme was conducted over three half days. The curriculum focused primarily on the identification of MR, which was defined as the presence of any retinal haemorrhages, retinal whitening or retinal vessel discolouration in the fundus. The curriculum also included identification of papilloedema, which is of prognostic significance in CM although alone not sufficient to define MR.^4^

The structure of the training program is outlined in Table 1.

| Session | Intended Learning Outcomes | Activities |
| --- | --- | --- |
| Day 1 | Knowledge acquisition<br>• Know the four signs of malarial retinopathy<br>• Recognise retinal haemorrhages in flat fundus photos<br>• Recognise retinal whitening in flat fundus photos<br>• Recognise retinal vessel change in flat fundus photos<br>• Recognise papilloedema in flat fundus photos | • Baseline assessment of knowledge of malarial retinopathy and papilloedema (20 fundus images)<br>• Didactic teaching on signs of malarial retinopathy<br>• Interactive practice identifying malarial retinopathy and papilloedema with immediate feedback (50 fundus images)<br>• Introduction to direct ophthalmoscopy |
| Day 3 | Skill acquisition<br>• Recognise papilloedema while using a direct ophthalmoscope in simulation eyes<br>• Identify Lea symbols in fundus using binocular indirect ophthalmoscopy | • Interactive practice identifying papilloedema in simulation eyes with direct ophthalmoscope with immediate feedback (10 eyes)<br>• Orientation to binocular indirect ophthalmoscopy<br>• Interactive practice identifying Lea symbols in simulation eyes with direct ophthalmoscope with immediate feedback (20 eyes)<br>• Assessment of ability to recognise |
|  |  | papilloedema in simulation eyes with direct ophthalmoscope (10 eyes) |
| Day 5 | Combine and consolidate<br>Recognise malarial retinopathy and papilloedema in simulation eyes using direct and binocular indirect ophthalmoscopy | Assessment of fundus examination for malarial retinopathy and papilloedema in simulation eyes (12 eyes) |

### Equipment

During the training, candidates used the Arclight BIO for indirect ophthalmoscopy. Direct ophthalmoscopy, primarily to visualise the optic nerve, was performed with the Arclight direct ophthalmoscope. Both have been developed with resource-limited settings in mind, being low-cost, solar-powered and robust. Both have been validated against much more expensive orthodox instruments and perform equally well.^7–9^ To simulate the retina we used a previously developed low-cost simulation eye.^10^ Model normal retinas with Lea symbols were printed onto matte paper using a high dots-per-inch inkjet printer. These were used to assess candidates’ ability to use the instrument in a way that was independent of clinical knowledge. An ophthalmologist with experience of MR (KJW) worked with a digital artist to design 12 model retinas with varying degrees of MR. Example images are shown in Figure 2. These simulation eyes were used to assess candidates’ ability to combine BIO with acquired clinical knowledge of MR.

**Figure 2.**
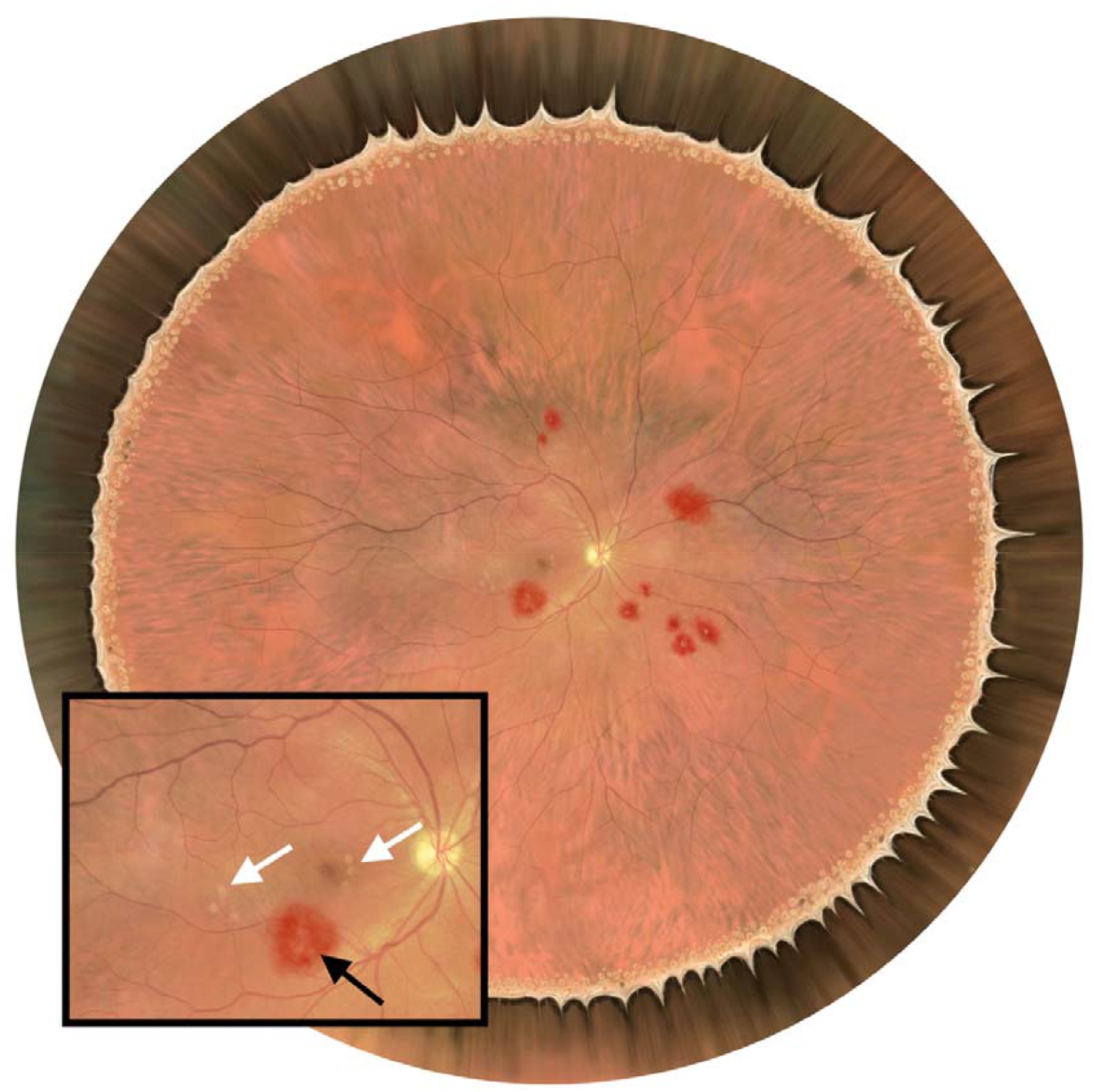
Simulated retina created by a digital artist and an ophthalmologist with experience of malarial retinopathy. Several haemorrhages and retinal whitening are visible. Inset: Zoomed in view of optic nerve and macula. A large, white-centred haemorrhage (black arrow) and a small amount of perifoveal and macula whitening are visible. There is no papilloedema.

### Pedagogical Methods

The training programme was designed according to the principles of Bloom’s taxonomy and constructive alignment.^11,12^ For practical tasks involving ophthalmoscopy we adopted Peyton’s four-step approach.^13^ This method promotes skill acquisition using a structured process comprising demonstration, deconstruction, comprehension and performance.

Briefly, we defined intended learning outcomes at the beginning of each session. We then conducted a baseline assessment, followed by a short didactic teaching session to transfer knowledge to the candidates. Subsequently, the candidates’ comprehension was assessed during practical activities. Following each task we conducted a post-activity assessment. Facilitators provided immediate feedback on candidates’ performance during the comprehension components of each activity.

### Statistics

Results of assessments were recorded anonymously on paper assessment forms during the training. A qualitative assessment of the candidates’ confidence with clinical assessment using direct and indirect ophthalmoscopy was completed online. The results were then input into Microsoft Excel v16.81 by the same examiner on two separate days.^14^ Discrepancies were checked against the paper forms and corrected. The data was then imported into R v4.3.2 for statistical analysis.^15^ Density plots and the Shapiro-Wilk test for normality were used to establish if the data were normally distributed. All data was paired and non-Gaussian so comparisons were performed using the Wilcoxon signed rank test with continuity correction.

To test the hypothesis that focussed simulation training would rapidly equip ophthalmology-naïve participants to identify malarial retinopathy, we compared the pre- and post-training scores between generalists (considered truly ophthalmology-naïve) and optometrists. Statisical testing for these two groups used the Wilcoxon rank-sum test for unpaired data.

## Results

### Quantitative analysis of training outcomes

The candidates’ performance in identifying malarial retinopathy by simply looking at fundal photographs prior to the training was mediocre, achieving a mean accuracy of 66%. Overall, candidates performed well at identifying papilloedema in photographs, even prior to training achieving a mean accuracy of 81%.

Following training, candidates were asked to identify MR using the BIO in simulation eyes. This test simultaneously assessed acquisition of knowledge and skill. The decision to combine the assessments was pragmatic; in a short training the burden of assessment can become onerous, and the delivery of high-quality program took precedence over data collection for the audit. Candidates had significantly improved their identification of MR following the training, achieving a mean accuracy of 83% (p = 0.014).

Similarly, the post-training assessment of papilloedema using the direct ophthalmoscope combined both knowledge and skill acquisition. Candidates’ knowledge had improved slightly after training achieving a mean accuracy of 85% (p = 0.30), though this did not reach statistical significance.

### Training outcomes by group

The participants were a mixture of generalists (paediatricians, nurses and general practitioners) and recently qualified optometrists. Optometrists had little to no experience of MR. It was assumed that optometrists had more experience with fundus photography and eye examination than generalists.

Before training, optometrists performed significantly better than generalists in identifying MR in flat images. Following training, there was no difference between the two groups when identifying MR using BIO. Results are presented in Table 2.

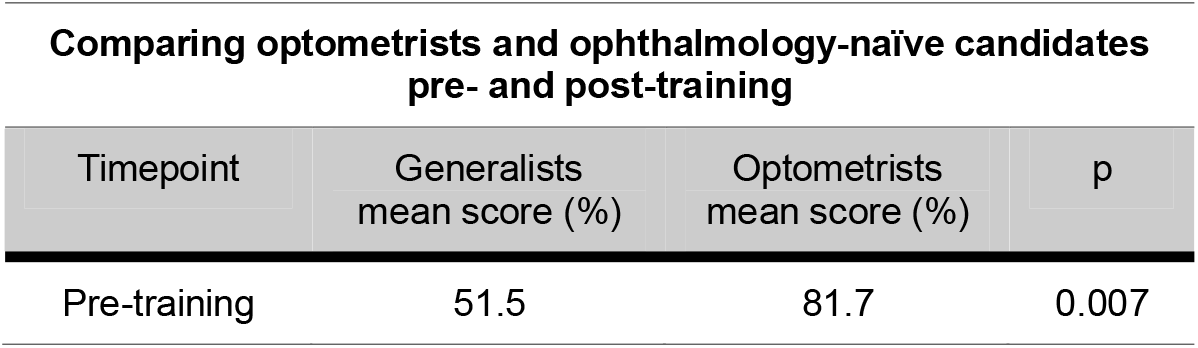
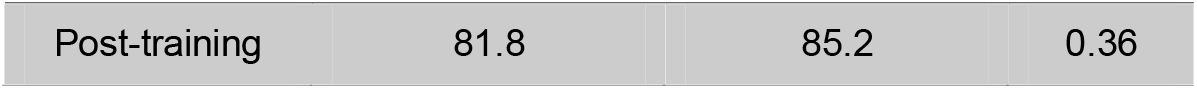

**Comparing optometrists and ophthalmology-naïve candidates pre- and post-training**
| Timepoint | Generalists mean score (%) | Optometrists mean score (%) | p |
| --- | --- | --- | --- |
| Pre-training | 51.5 | 81.7 | 0.007 |
| Post-training | 81.8 | 85.2 | 0.36 |

### Qualitative analysis of training outcomes

Course participants were asked to give feedback on the course. Specifically, they were asked the following questions both before and after the training:

1. How confident are you that you would recognise the signs of malarial retinopathy in a flat image?
2. How confident are you that you would recognise the signs of malarial retinopathy in an eye?
3. How confident are you at identifying papilloedema with a direct ophthalmoscope?
4. How confident are you at identifying retinal haemorrhages with an indirect ophthalmoscope?
5. How confident are you at identifying retinal whitening with an indirect ophthalmoscope?
6. How confident are you at identifying retinal vessel change with an indirect ophthalmoscope?

Each question was scored using a 7-point Likert scale where 1 represents very low confidence and 7 represents very high confidence.

Perceived confidence increased in all domains. The results are summarised in Table 3.

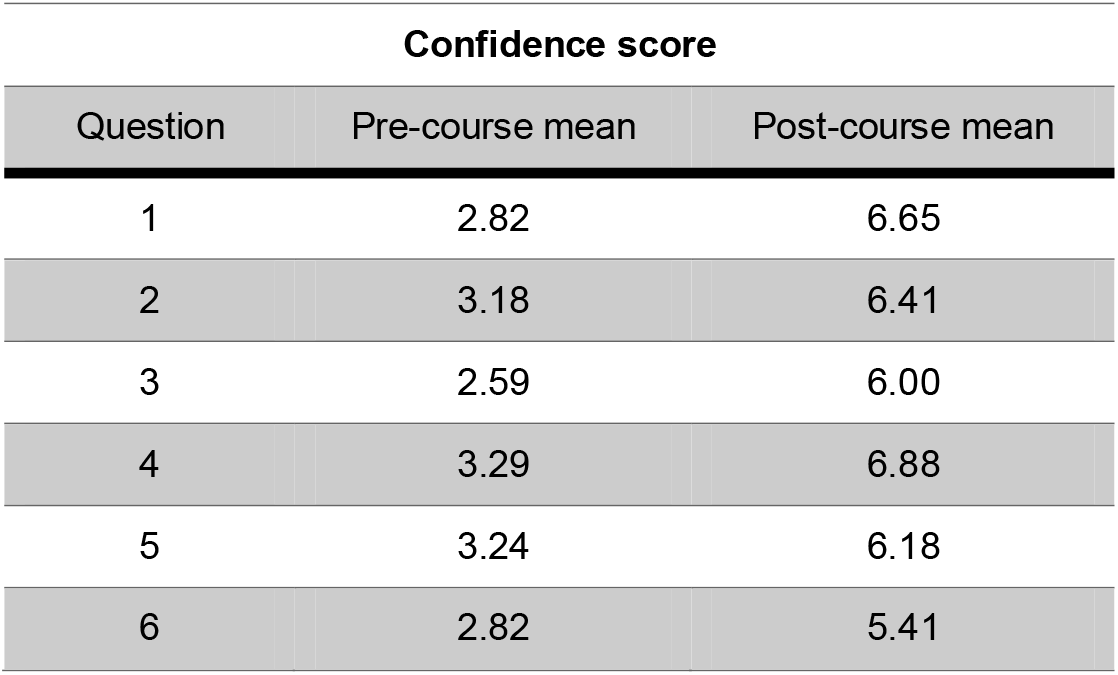

## Discussion

The findings of this audit demonstrate the feasibility and effectiveness of a targeted training program aimed at non-ophthalmologist healthcare providers for the identification of MR using low-cost training tools and ophthalmic equipment. Given the typically limited access to specialized ophthalmic care in regions with a high burden of CM, the ability to rapidly and accurately diagnose MR has the potential to improve diagnosis and management.^6^

These results demonstrate a clear improvement in candidates’ ability to identify MR following the structured simulation training program. Prior to the training, participants exhibited only moderate proficiency in identifying MR in flat images. After the training they were able to diagnose MR with a high level of accuracy. This improvement is particularly significant considering that during the post-training assessment candidates were using a BIO, highlighting the potential for non-ophthalmologist healthcare providers to quickly acquire proficiency in a more sophisticated form of wide-field binocular ophthalmoscopy typically considered the preserve of eye specialists. Moreover, ophthalmology-naïve practitioners attained a similar level of proficiency to optometrists immediately following the training. This dispels the widely held view that binocular indirect ophthalmoscopy is a skill that specialists can acquire only after a long period of training and practice. This finding demonstrates that this form of ophthalmoscopy, for specific use-cases, can therefore be learned rapidly and to a high level by ophthalmology-naïve generalists for scaling in more general clinical settings.

In this study the integration of practical, hands-on training sessions using simulation eyes enabled participants to apply theoretical knowledge to simulated real-world scenarios. Simulation is well-established in medical education and has been successfully employed in ophthalmology in various settings.^16^ The simulation eyes designed for this program provide an inexpensive yet valuable resource for novice users to practice and refine their skills in a controlled environment, without the need for live patient encounters or expensive virtual reality equipment. In addition to the education benefits, simulation reduces risk to sick patients by minimising lengthy, potentially uncomfortable, examinations by multiple inexperienced users while training.

The observed increase in participants’ confidence levels across various domains of retinal examination further supports the efficacy of the training program. Confidence is a key determinant of clinical competence, and the positive shift in participants’ confidence levels suggests that they will be better equipped to identify MR in clinical practice.^17^

The strength of the training program is its emphasis on combining knowledge with skill acquisition using a structured pedagogical approach. Sequentially guiding participants through didactic teaching, reviewing their comprehension, and demonstrating and practicing skills with timely feedback facilitated the transfer of knowledge into clinically relevant skill acquisition. This systematic approach is grounded in educational theory and can serve as a model for future training initiatives aimed at enhancing diagnostic capabilities at scale.

It should be noted that the short-term nature of the training program limits the ability to assess long-term retention of skills beyond the immediate post-training period. Future studies should incorporate longitudinal follow-up to evaluate the sustainability of skill acquisition over time in real world clinical settings.

## Conclusions

MR has diagnostic significance in regions where asymptomatic malarial parasitaemia can cast doubt on the major contributing cause of coma in febrile children who meet the WHO criteria for CM. Structured simulation-based training for non-ophthalmologists can rapidly equip frontline healthcare workers in malaria-endemic areas with the necessary skills to diagnose malarial retinopathy in simulation eyes. Further work to assess the retention of skills beyond the immediate post-training period is required.

## Supporting information

Supplementary Information: Reproducible Analysis

## Data Availability

All data produced in the present study are available upon reasonable request to the authors.

## Ethics approval and consent to participate

The requirement for formal ethical approval and participant consent for this study was waived by the College of Medicine Research Ethics Committee, Kamuzu University of Health Sciences, Malawi. COMREC Reference Number P.10/24-1186.

## Consent for publication

All individuals named in this manuscript have given their consent to publish.

## Availability of data and material

For training materials, please contact The Arclight Project at the Univeristy of St. Andrew’s. The data required to reproduce this analysis is available as supplementary information.

## Competing interests

The authors declare no competing interests.

## Funding

This study was funded by the Wellcome Institutional Translation Partnership Award at Malawi-Liverpool-Wellcome Trust (Grant ID: 219633/Z/19/Z).

## Authors’ contributions

KJW, OK & AB conceived the idea and developed simulation retinas with the artist; KJW secured funding, conducted the analysis and drafted the manuscript; KJW, OK, HN & AL designed and delivered the training; NAVB provided expert input on malarial retinopathy and provided direct supervision to KJW; all authors reviewed, edited and approved the final manuscript.

## Acknowledgements

We, the authors, would like to acknowledge the significant work undertaken by artists Matthew Briggs (UK) and Clement Kammwamba (Malawi). For details on how to contact either artist for medical illustration, please contact the corresponding author. Equally, we would like to thank all the participants at the ‘Fundoscopy for Cerebral Malaria’ course.

## Notes

### Competing Interest Statement

The authors have declared no competing interest.

### Author Declarations

Ethics committee of Kamuzu University of Health Sciences waived ethical approval for this work.

