## Supplementary Information: Reproducible Analysis for "Structured simulation-based training with frugal ophthalmic instruments rapidly equips non-ophthalmologists to identify malarial retinopathy": 20231116_BIOCM_DataAnalysis_v2.0.pdf

The results of all quantitative tests were collected on paper forms during the course. The answers were input into a Microsoft Excel spreadsheet by the same grader on two separate days. Discrepancies were checked by the same grader and corrected where necessary. The spreadsheet automatically checked the candidates answers against a refence and calculated the percentage score for each assignment. This data was then imported into R.

The results of the qualitative tests were collected in Google forms and exported into Microsoft Excel. After formatting and manual validation, the data was imported into R.

A column was added to each of the imported data frames to indicate whether data was complete for that candidate. Individuals who has missing data were excluded from downstream analyses. To avoid mistaking the candidate number with numerical data, each value was prepended with "bio".

```
# import data from quantitative tests done during the course
test <- read_xlsx(here("data/20231110_BIOCM_Analysis_v1.0.xlsx"), sheet = 5)

# import data from qualitative surveys pre- and post-course
qual <- read_xlsx(here("data/20231110_BIOCM_Analysis_v1.0.xlsx"), sheet = 6)

# add a column to each df to indicate if data is complete
test$complete <- apply(test, 1, function(row) ifelse(any(is.na(row)), 0, 1))
qual$complete <- apply(qual, 1, function(row) ifelse(any(is.na(row)), 0, 1))
```

```r
# add "bio" to candidate numbers to avoid mistaking for numerical data
test$cand_no <- paste0("bio", test$cand_no)
qual$cand_no <- paste0("bio", qual$cand_no)

# add group variable
test$group <- c("general", "general", "general", "general", "general",
                "general", "general", "general", "general", "optom",
                "optom", "general", "optom", "optom", "optom",
                "optom", "optom", "optom", "optom", "general")
```

### Quantitative analysis

We want to test the hypothesis that following training, the candidates are as good or better at detecting malarial retinopathy and papilloedema using ophthalmoscopes as they were before the course using flat images.

To do this, we perform a two-sided Mann-Whitney-U test on the before and after data for malarial retinopathy and papilloedema. If the value for the difference in the means is positive, this confirms an improvement in scores following training.

```r
# check for normality
# ggdensity(test$mr_bio_post_pc)
# ggqqplot(test$mr_bio_post_pc)
# shapiro.test(test$mr_bio_post_pc)

# ggdensity(test$mr_flat_pre_pc)
# ggqqplot(test$mr_flat_pre_pc)
# shapiro.test(test$mr_flat_pre_pc)

# perform Mann-Whitney-U test for malarial retinopathy (non-Gaussian)
mr_result <- test %>%
  filter(complete == 1) %>%
  with(wilcox.test(mr_bio_post_pc, mr_flat_pre_pc,
             alternative = "two.sided", paired = TRUE))
```

```
Warning in wilcox.test.default(mr_bio_post_pc, mr_flat_pre_pc, alternative =
"two.sided", : cannot compute exact p-value with ties
```

```r
# check for normality
# ggdensity(test$pap_arc_post_pc)
# ggqqplot(test$pap_arc_post_pc)
```

```r
# shapiro.test(test$pap_arc_post_pc)

# ggdensity(test$pap_flat_pre_pc)
# ggqqplot(test$pap_flat_pre_pc)
# shapiro.test(test$pap_flat_pre_pc)

# perform Mann-Whitney-U test for papilloedema (non-Gaussian)
pap_result <- test %>%
  filter(complete == 1) %>%
  with(wilcox.test(pap_arc_post_pc, pap_flat_pre_pc,
              alternative = "two.sided", paired = TRUE))
```

```
Warning in wilcox.test.default(pap_arc_post_pc, pap_flat_pre_pc, alternative =
"two.sided", : cannot compute exact p-value with ties
```

```
Warning in wilcox.test.default(pap_arc_post_pc, pap_flat_pre_pc, alternative =
"two.sided", : cannot compute exact p-value with zeroes
```

```r
mr_result
```

```

    Wilcoxon signed rank test with continuity correction

data:  mr_bio_post_pc and mr_flat_pre_pc
V = 156.5, p-value = 0.01404
alternative hypothesis: true location shift is not equal to 0
```

```r
pap_result
```

```

    Wilcoxon signed rank test with continuity correction

data:  pap_arc_post_pc and pap_flat_pre_pc
V = 99, p-value = 0.2958
alternative hypothesis: true location shift is not equal to 0
```

Here, our data suggest that candidates were better at identifying malarial retinopathy in simulation eyes with an indirect ophthalmoscope after the course than they were at detecting malarial retinopathy in flat images prior to the course. This demonstrates acquisition of knowledge about the signs of the pathology, and acquisition of skill using the indirect ophthalmoscope.

There was no observable improvement in the detection of papilloedema, but it should be noted that candidates were highly capable of detecting papilloedema in flat images before the course. This demonstrates that following the course, candidates were able to reliably detect papilloedema in simulation eyes using direct fundoscopy.

To check whether previous eye-specific training experience affected attainment during the course, we compared the test performance between early-career optometrists and generalists.

```
by_group <- test %>%
  group_by(group) %>%
  summarise(
    mean_pre = mean(mr_flat_pre_pc, na.rm = T),
    mean_post = mean(mr_bio_post_pc, na.rm = T)
)

# Wilcoxon rank-sum test for pre values
wilcox_test_pre <- wilcox.test(
  mr_flat_pre_pc ~ group,
  data = test,
  exact = FALSE
)

# Wilcoxon rank-sum test for post values
wilcox_test_post <- wilcox.test(
  mr_bio_post_pc ~ group,
  data = test,
  exact = FALSE
)

# print results
by_group
```

```
# A tibble: 2 x 3
  group   mean_pre mean_post
  <chr>      <dbl>     <dbl>
1 general     51.5      81.8
2 optom       81.7      85.2
```

```
wilcox_test_pre
```

```

	Wilcoxon rank sum test with continuity correction

data:  mr_flat_pre_pc by group
W = 11.5, p-value = 0.006512
alternative hypothesis: true location shift is not equal to 0
```

```
wilcox_test_post
```

```

	Wilcoxon rank sum test with continuity correction

data:  mr_bio_post_pc by group
W = 37.5, p-value = 0.3636
alternative hypothesis: true location shift is not equal to 0
```

### Qualitative analysis

Candidates were asked a series of questions relating to their confidence identifying malarial retinopathy in images and in eyes both before and after the course. Answers were given using a 7-point Likert scale. The questions have been summarised below:

1) How confident are you that you would recognise the signs of malarial retinopathy in a flat image?
2) How confident are you that you would recognise the signs of malarial retinopathy in an eye?
3) How confident are you that you could identify papilloedema using direct ophthalmoscopy?
4) How confident are you that you could identify retinal haemorrhages using indirect ophthalmoscopy?
5) How confident are you that you could identify retinal whitening using indirect ophthalmoscopy?
6) How confident are you that you could identify retinal vessel change using indirect ophthalmoscopy?

To demonstrate that previously inexperienced clinicians gained confidence in the identification of malarial retinopathy and papilloedema using ophthalmoscopy, we present the mean scores pre- and post-training for each of the questions for all participants who completed both surveys (n = 17).

```r
flat <- qual %>%
  filter(complete == 1) %>%
  summarise(
    flat_pre = mean(conf_flat_pre),
    flat_post = mean(conf_flat_post),
    flat_diff = t.test(conf_flat_post, conf_flat_pre,
                       alternative = "two.sided", paired = TRUE)$estimate,
    flat_lci = t.test(conf_flat_post, conf_flat_pre,
                       alternative = "two.sided", paired = TRUE)$conf.int[1],
    flat_uci = t.test(conf_flat_post, conf_flat_pre,
                       alternative = "two.sided", paired = TRUE)$conf.int[2],
    flat_p = t.test(conf_flat_post, conf_flat_pre,
                       alternative = "two.sided", paired = TRUE)$p.value)

eye <- qual %>%
  filter(complete == 1) %>%
  summarise(
    eye_pre = mean(conf_eye_pre),
    eye_post = mean(conf_eye_post),
    eye_diff = t.test(conf_eye_post, conf_eye_pre,
                       alternative = "two.sided", paired = TRUE)$estimate,
    eye_lci = t.test(conf_eye_post, conf_eye_pre,
                       alternative = "two.sided", paired = TRUE)$conf.int[1],
    eye_uci = t.test(conf_eye_post, conf_eye_pre,
                       alternative = "two.sided", paired = TRUE)$conf.int[2],
    eye_p = t.test(conf_eye_post, conf_eye_pre,
                       alternative = "two.sided", paired = TRUE)$p.value)

pap <- qual %>%
  filter(complete == 1) %>%
  summarise(
    pap_pre = mean(conf_pap_pre),
    pap_post = mean(conf_pap_post),
    pap_diff = t.test(conf_pap_post, conf_pap_pre,
                       alternative = "two.sided", paired = TRUE)$estimate,
    pap_lci = t.test(conf_pap_post, conf_pap_pre,
                       alternative = "two.sided", paired = TRUE)$conf.int[1],
    pap_uci = t.test(conf_pap_post, conf_pap_pre,
                       alternative = "two.sided", paired = TRUE)$conf.int[2],
    pap_p = t.test(conf_pap_post, conf_pap_pre,
                       alternative = "two.sided", paired = TRUE)$p.value)
```

```
haem <- qual %>%
  filter(complete == 1) %>%
  summarise(
    haem_pre = mean(conf_haem_pre),
    haem_post = mean(conf_haem_post),
    haem_diff = t.test(conf_haem_post, conf_haem_pre,
                       alternative = "two.sided", paired = TRUE)$estimate,
    haem_lci = t.test(conf_haem_post, conf_haem_pre,
                       alternative = "two.sided", paired = TRUE)$conf.int[1],
    haem_uci = t.test(conf_haem_post, conf_haem_pre,
                       alternative = "two.sided", paired = TRUE)$conf.int[2],
    haem_p = t.test(conf_haem_post, conf_haem_pre,
                       alternative = "two.sided", paired = TRUE)$p.value)

white <- qual %>%
  filter(complete == 1) %>%
  summarise(
    white_pre = mean(conf_white_pre),
    white_post = mean(conf_white_post),
    white_diff = t.test(conf_white_post, conf_white_pre,
                       alternative = "two.sided", paired = TRUE)$estimate,
    white_lci = t.test(conf_white_post, conf_white_pre,
                       alternative = "two.sided", paired = TRUE)$conf.int[1],
    white_uci = t.test(conf_white_post, conf_white_pre,
                       alternative = "two.sided", paired = TRUE)$conf.int[2],
    white_p = t.test(conf_white_post, conf_white_pre,
                       alternative = "two.sided", paired = TRUE)$p.value)

vc <- qual %>%
  filter(complete == 1) %>%
  summarise(
    vc_pre = mean(conf_vc_pre),
    vc_post = mean(conf_vc_post),
    vc_diff = t.test(conf_vc_post, conf_vc_pre,
                       alternative = "two.sided", paired = TRUE)$estimate,
    vc_lci = t.test(conf_vc_post, conf_vc_pre,
                       alternative = "two.sided", paired = TRUE)$conf.int[1],
    vc_uci = t.test(conf_vc_post, conf_vc_pre,
                       alternative = "two.sided", paired = TRUE)$conf.int[2],
    vc_p = t.test(conf_vc_post, conf_vc_pre,
                       alternative = "two.sided", paired = TRUE)$p.value)
```

```r
# PLEASE NOTE THAT DUE TO NON-NORMALITY OF THE DATA CONFIDENCE INTERVALS AND
# P-VALUES HAVE BEEN EXCLUDED FROM THE FINAL DATA PRESENTATION

header <- c("pre", "post", "diff", "lci", "uci", "p")
colnames(flat) <- header
colnames(eye) <- header
colnames(pap) <- header
colnames(haem) <- header
colnames(white) <- header
colnames(vc) <- header
question <- 1:6

tab <- as.data.frame(rbind(flat, eye, pap, haem, white, vc))
tab <- tab %>%
  mutate(across(1:5, ~round(., 2))) %>%
  mutate(p = ifelse(p < 0.001, "<0.001", round(p, 3)))
tab <- tab %>%
  transmute(
    pre = pre,
    post = post
  )
tab <- cbind(question, tab)

ft <- flextable(tab) %>%
  set_header_labels(question = "Question", pre = "Pre-course mean",
                    post = "Post-course mean") %>%
  add_header_row(values = c("","Confidence score"),
                 colwidths = c(1, 2)) %>%
  align(align = "center", part = "all") %>%
  autofit()

ft
```

| | Confidence score | |
|---|---|---|
| Question | Pre-course mean | Post-course mean |
| 1 | 2.82 | 6.65 |
| 2 | 3.18 | 6.41 |
| 3 | 2.59 | 6.00 |
| 4 | 3.29 | 6.88 |

| | Confidence score | |
|---|---|---|
| Question | Pre-course mean | Post-course mean |
| 5 | 3.24 | 6.18 |
| 6 | 2.82 | 5.41 |
